# Online early supportive psychological intervention for bereaved families during COVID-19: Lessons Learned and Implementation Challenges

**DOI:** 10.1101/2024.02.03.24302281

**Authors:** Mona Hashemzadeh, Samira Jamaloo, Elnaz Shahmohamadi, Anahita Rahmani, Gilda Rajabi Damavandi, Mehri Seyed Mousavi, Hossein Gharaati Sotoudeh, Valentin Artounian

## Abstract

As the COVID-19 pandemic has created complex conditions and the horrific loss of numerous lives, grieving the loss of loved ones in close families can be extremely difficult. To reduce the suffering of the loss and prevent the development of complicated grief, it is necessary to provide bereavement care as soon as possible. Therefore, we quickly developed a complete online program that included supportive psychotherapeutic interventions and psychiatric counseling. The structure of all services is the main emphasis of the study, which also emphasizes the quantitative components and the unique characteristics of the interventions. Based on the lesson learned, we discussed the difficulties experienced in putting into practice an internet-based preventive service.

## 1. Introduction

The emergence of the 2019 coronavirus (COVID-19) in late December 2019 in Wuhan, China, was the beginning of a global health crisis. In response to the worsening situation, the World Health Organization (WHO) declared the outbreak of COVID-19 an international public health emergency. As of the end of January, 2024, more than seven million people worldwide, including 146,800 Iranians, have died from the disease (World Health Organization, 2024), leading to widespread grief among those affected (Zhai and Du, 2020). Unfortunately, social isolation and frequent quarantines have become the primary preventive measures against COVID-19 due to the lack of specific medical treatments. While the goal of these isolation strategies is to contain the spread of the virus, they inadvertently reduce access to family and social support systems and increase feelings of loneliness, anxiety, and depression (Shah et al., 2021). Additionally, restrictions on gatherings, including funerals and related religious and cultural rituals, have deprived families of vital social support when they need it most, potentially exacerbating the bereavement experience.

Loss and grief are natural aspects of human life, often resolved spontaneously over time (Schwartz-Borden, 1986). However, approximately 10-15% of individuals may experience prolonged and profound feelings of grief, with about 9% meeting the criteria for complicated grief (Currier et al., 2008; Wittouck et al., 2011). Additionally, recently bereaved individuals face an elevated risk of developing other psychiatric disorders, emphasizing the need for effective preventive interventions (Mayland et al., 2020; Yalom and Vinogradov, 1988).

Research has demonstrated the crucial role of social support in moderating grief outcomes during the grieving process (Neimeyer et al., 2014). Specifically, enhancing social support that focuses on emotional aspects may help alleviate the loneliness experienced by grievers during the COVID-19 pandemic. Emotional support in grief entails being present, spending time with the grieving individual, listening to their narratives without judgment, and acknowledging the deceased loved one through reminiscing and using their name. It also involves refraining from imposing time limits on the grieving process (Cacciatore et al., 2021). Based on this understanding, therapists have developed various supportive interventions, delivered in individual or group formats, to prevent and treat grief (Hoffmann et al., 2018; Kang and Yoo, 2007; Nam, 2016). Considering the limitations imposed by COVID-19 quarantine measures, group interventions appear more feasible as they provide a space for sharing emotions, receiving emotional support and validation, normalizing the grieving experience through universalization, gaining knowledge about the grieving process, learning coping techniques from others, and fostering a sense of belonging to a community, thus mitigating isolation (DeLucia-Waack et al., 2014; Steiner, 2006; Yopp and Rosenstein, 2013). However, challenges such as late arrival or non-continuation of participation by some members and the short duration of these groups can evoke grief-related emotions and necessitate the development of new coping strategies (Marmarosh et al., 2020).

Despite the existing studies on bereavement support groups, several review articles have found no significant impact of preventive interventions on grief outcomes. Currier et al. (2008) reported that grief interventions were effective primarily for individuals who experienced difficulties adapting to grief or exhibited symptoms of psychiatric disorders. Another meta-analysis by Wittouck et al. (2011) indicated that preventive measures had limited effects on clients, while treatment interventions demonstrated effectiveness in both short-term and long-term follow-ups. The authors suggested that individuals experiencing grief, whether their reaction is considered normal or complicated, would benefit from therapy when they perceive the need for counseling. It has been suggested that evaluations of these treatments should be qualitative rather than quantitative and concentrate on the support given (Wittouck et al., 2011). The results were more promising in the latest review article by M. Johannsen et al. (2019) about grief. It has been shown that psychological interventions related to grief in adults significantly affect grief symptoms over time, specifically found that interventions would be more effective if delivered within the first six months of loss and for people with significant grief symptoms. In contrast to previous studies suggesting similar effects of group and individual interventions on grief, this study indicated that individual interventions were more effective, although caution should be exercised when interpreting the results.

Additionally, the utilization of online treatment methods has grown significantly in recent years. Several studies have demonstrated the efficacy of online therapy, which is comparable to face-to-face therapy (Wagner et al., 2014). Online support groups have also emerged as reliable sources of help, capable of improving the well-being of grieving individuals. For instance, a review study on online grief groups conducted by Robinson and Pond (2019) observed that when the therapeutic effects were examined qualitatively, participants derived therapeutic benefits from interventions, including emotional support, a sense of community, and information sharing. Also, online studies on grief prevention have been planned for individual therapy, including Hoffman et al.’s study (2018). It is assumed that these online interventions can potentially yield beneficial effects for individuals experiencing grief. Children and teenagers, like adults, experience grief following a loss; however, their reactions may vary based on their age and level of development. Currier et al.’s meta-analysis (2008) found that interventions for grief in children exhibited similar effectiveness to those in adults.

During the COVID-19 outbreak, literature has proposed various strategies to support grieving families and prevent pathological grief. Particularly, the importance of maintaining follow-up contact with bereaved relatives, including through phone calls, has been emphasized (Carr et al., 2020; Selman et al., 2020). For example, in a study conducted by Menichetti Delor et al. (2021) psychologists-initiated contact with the families of COVID-19 patients who had passed away two to three days after the death occurred in the hospital. These contacts provided a supportive environment for expressing grief-related emotions, mobilizing internal psycho-emotional resources, and making referrals for more severe symptoms. Additionally, Dominguez-Rodriguez et al. (2021) launched an online service in Mexico to enhance the quality of life during the COVID-19 outbreak with offering self-applied cognitive behavioral therapy (CBT) interventions.

Addressing the emotional impact of grief, creating a supportive environment, raising awareness about the grief process, and strengthening coping mechanisms can help alleviate grief symptoms. Thus, this study aims to report our experience in establishing an online treatment service in Iran for bereaved individuals who have lost an immediate family member during the COVID-19 outbreak. The goal is to prevent complicated grief and contribute to the current body of knowledge on effective interventions for grief during the pandemic.

## 2. Methods

### 2.1. Setting

We obtained a list of individuals who died due to COVID-19 between March 2020 and September 2020 from three central hospitals affiliated with Tehran University of Medical Sciences (Ziaeian, Baharlu, and Imam Khomeini). As proposed by Menichetti Delor et al. (2021) in our study, skilled clinical psychologists initiated contact with immediate family members of the deceased who had lived with the patient, assessed their psychological states, and introduced psychotherapeutic treatments. In cases where the hospital did not have a collaborating psychologist, contacts were made by collaborating psychologists from Roozbeh Hospital, the main psychiatric hospital of Tehran University.

### 2.2. Intervention protocol

#### 2.2.1. Screening

As shown in Figure 1, in the first step, psychologists screened first-degree family members based on the severity of psychological impairment due to the loss, identifying those requiring urgent psychiatric referral. In the second step, the residual survivors who were not severely psychologically impaired but had not yet coped with their loss offered supportive psychotherapy. If a bereaved person referred another immediate family member for screening, the psychologist contacted the individual and repeated the screening process. For participants under 18 years of age, the same project psychologists conducted the initial screening.

**Figure 1.**
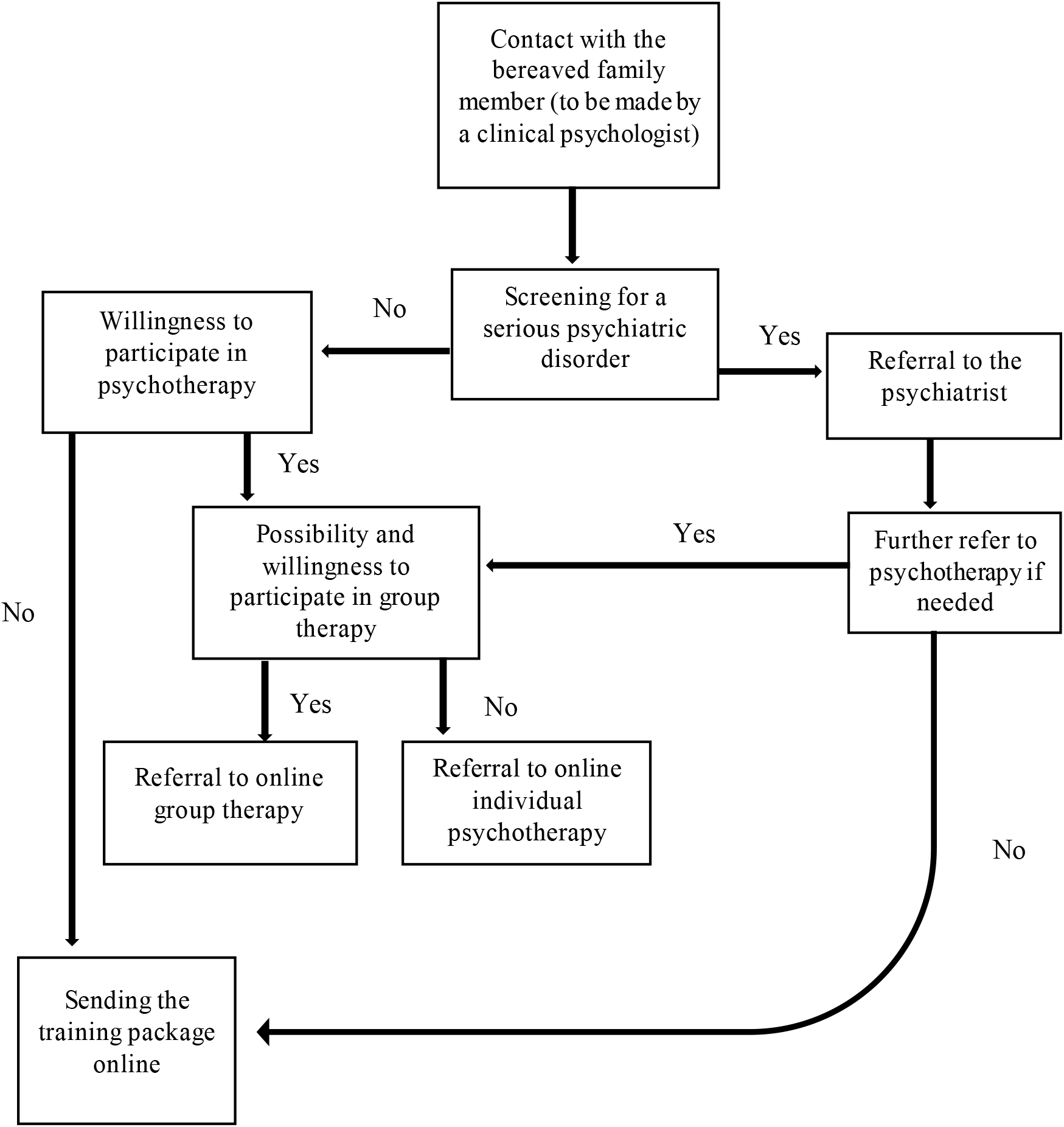
The intervention protocol

At the end of the screening phone call, we sent educational materials on grief to all participants. Referrals were based on our service algorithm and psychologists’ clinical judgment. Since each psychologist has a different approach to working with clients, we designed a unique algorithm and communication guide for the initial contacts to ensure consistency among psychologists (see Table 1).

**Table 1.**
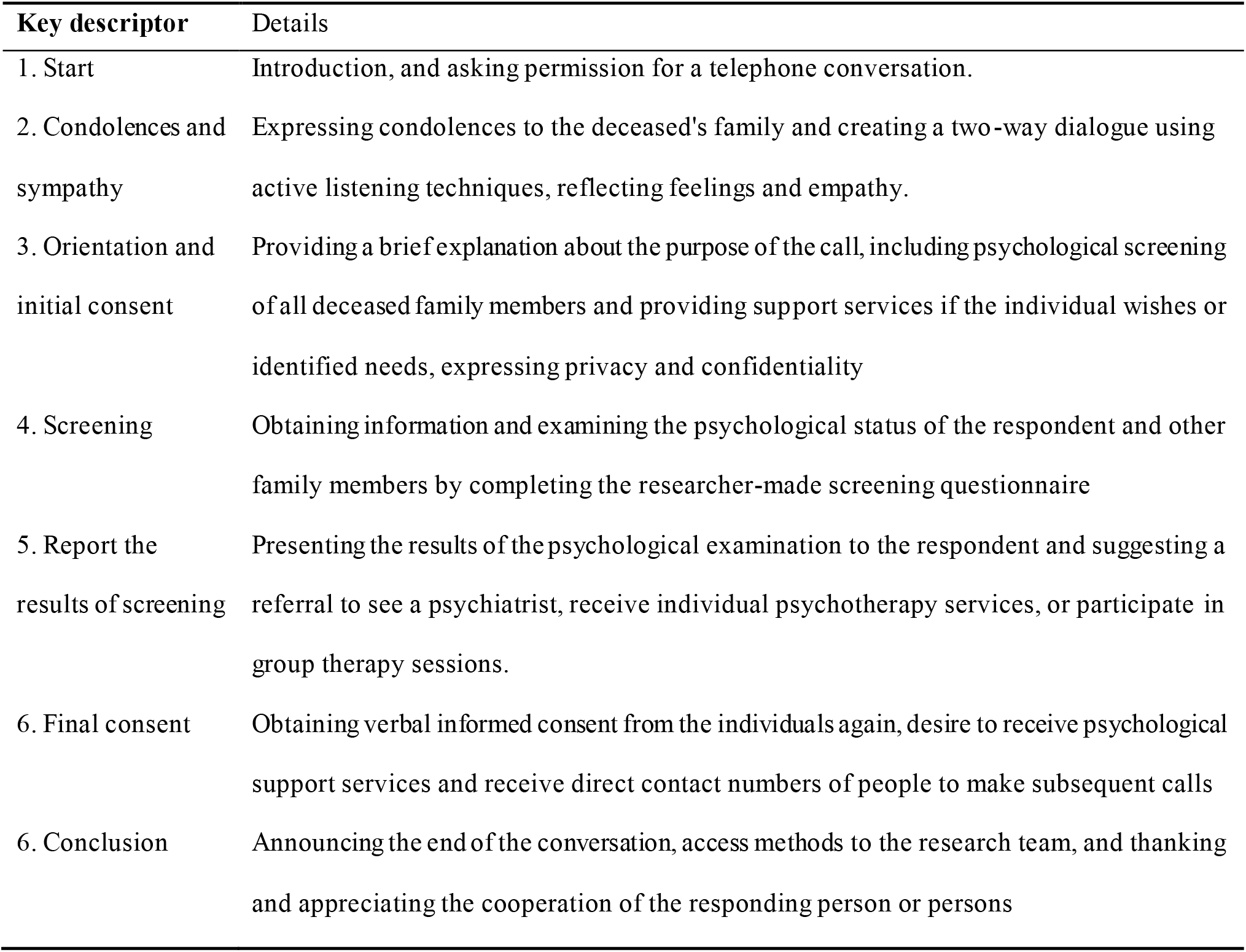
The content of phone calls with the bereaved family manual.

#### 2.2.2. Psychiatric Consultation

Symptoms of severe psychological impairments included mood swings, irritability, restlessness, severe somatic symptoms, thoughts of self-harm or aggression toward others, suicidal behaviors, vegetative symptoms such as anorexia or severe insomnia, denial of loss, functional impairment, recurrence of previous psychiatric symptoms, and the need for revising psychiatric medication regimens. After active screening, we recommended psychiatric visits for these individuals. Within a few days, participants who consented were scheduled for audio or video online psychiatric visits. These visits were facilitated by second-or third-year psychiatric residents, with supervision provided by a faculty member involved in the study. For children and adolescents, the same procedure was followed, and those with severe psychological impairments were referred to a child psychiatrist for online visits.

Following psychiatric visits, participants were introduced to supportive psychotherapy interventions once their severe symptoms had subsided, as determined by the clinical judgment of the psychiatry residents.

#### 2.2.3. Supportive Psychotherapy

The supportive psychotherapy intervention program included online group therapy and online/phone call individual psychotherapy. All participants were first referred to online supportive group therapy. Those who required one-on-one sessions or experienced technological issues were referred to individual supportive psychotherapy. To ensure consistency among therapists, a grief guideline was prepared, outlining therapeutic goals such as getting out of denial and accepting grief, understanding emotional reactions, discussing feelings of sadness, anger, and guilt, reviewing life memories with the deceased, and establishing new relationships and behaviors(Schwartz-Borden, 1986). Finally, the dos and do nots when working with clients were considered (see supplementary table 1).

##### A: Online Supportive Group Therapy

This approach primarily provided supportive, short-term, single-gender online group therapy for bereavement. Based on supportive psychotherapy, these group treatments addressed the difficulties associated with loss and the symptoms of grieving. Through active engagement with participants and a focus on conscious processes, the therapist facilitated the process. Non-analytical interventions were provided, and patients were encouraged to discuss their experiences and adaptation. Positive strategies were reinforced, and patients contributed to problem-solving techniques. The therapist also provided the necessary education and allowed patients to express their thoughts, ideas, and personal information.

Eight one-hour sessions per week, with a maximum of eight participants comprised the groups. Skype was used for the sessions, and prior to the session, participation policies and procedures were distributed (see Supplementary Table 2). Two therapists worked with each group: a psychiatrist served as the primary therapist and a third-year psychiatric resident served as the co-therapist. After each session, the two therapists discussed the session’s content and interventions. In cases where any group member required additional interventions, individual sessions were conducted by the same group psychotherapist to address their specific needs.

##### B: Online Individual Supportive Psychotherapy

The first stage in putting individual supportive psychotherapy into practice was to choose third-year residents in psychiatry who offered to serve as the individual’s therapists. These residents completed a thorough training program that included seven 90-minute sessions covering grief counseling guidelines and supportive psychotherapy techniques (Winston et al., 2020; Worden, 2018). After training, the residents started working along with having online supervision, which helped them conduct sessions more efficiently. One to eight sessions of individual supportive psychotherapy were held weekly, either online or through phone calls. The intervention was conducted using modified versions of the group principles and guidelines.

For participants below 18, an experienced child and adolescent psychologist provided individual supportive psychotherapy. The intervention was conducted online or via phone calls with the child or adolescent and their family members. The number of sessions was determined based on the needs of each child and their family. These sessions were designed to support children and adolescents as they grieved and to provide psycho-education for family members.

## 3. Results

### 3.1. Screening

A total of 605 deceased patients were recruited from selected COVID-19 tertiary hospitals between March 2020 and September 2020. Following the discussed method, we initiated contact with the immediate family of the departed through telephone communication. However, 116 calls (19.17%) were left unanswered (after two attempts). Out of 489 contacted families, 558 first-degree relatives were screened and entered the study.

### 3.2. Psychiatry Consultation and Percentage of Pre-treatment Dropout

Out of the total screened participants, 31.72% (n=177) needed a psychiatric consultation, as shown in Figure 2. Within this group, a psychiatric resident met with 33 women, nine men, and three children and adolescents. The remaining individuals declined consultations and were classified as pre-treatment dropouts, as they dropped out of the study before the commencement of psychiatric visits or supportive psychotherapy. According to the data, 14 cases (33.33%) of the adults who received psychiatric consultation were diagnosed with normal grief. The remaining adults, as well as all children and adolescents, were diagnosed with depression and/or anxiety and were prescribed medication.

**Figure 2.**
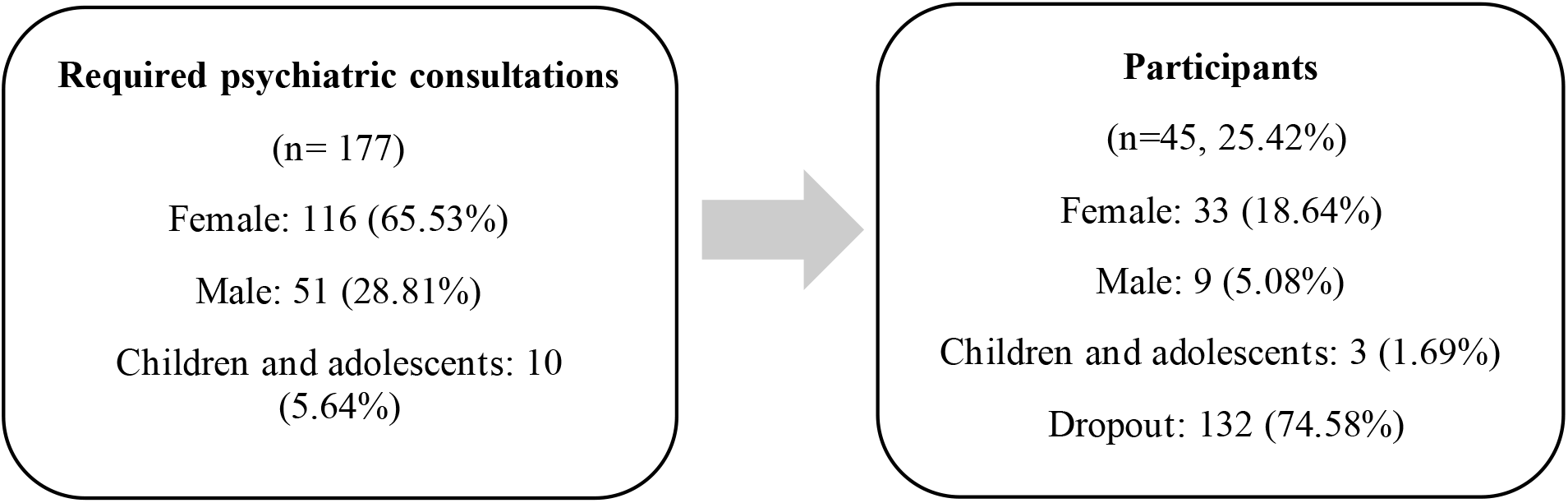
The percentage of dropouts in the group required psychiatric visits

### 3.3. Supportive Psychotherapy and Percentage of Pre-treatment Dropout

Among the screened individuals, 35.12% (n=196) expressed their willingness to partake in supportive psychotherapy (Figure 3). The main aim was to create single-sex groups for online supportive group therapy, the primary intervention for adults. Despite all the efforts researchers made, various participants preferred individual supportive psychotherapy over group therapy. Consequently, a modification was made to the psychotherapy referral process amid the project. As a result, all adults who required psychotherapy were referred to individual supportive psychotherapy. Participation in psychotherapy sessions had a significant pre-treatment dropout rate (80.61%), as shown in Figure 3. Out of the women who wanted supportive psychotherapy, 22.80% took part in the sessions, compared to only 7.35% of men. In the group of children and adolescents, the pre-treatment dropout rate was 50.0%, and more girls than boys (six girls vs. one boy) participated.

**Figure 3.**
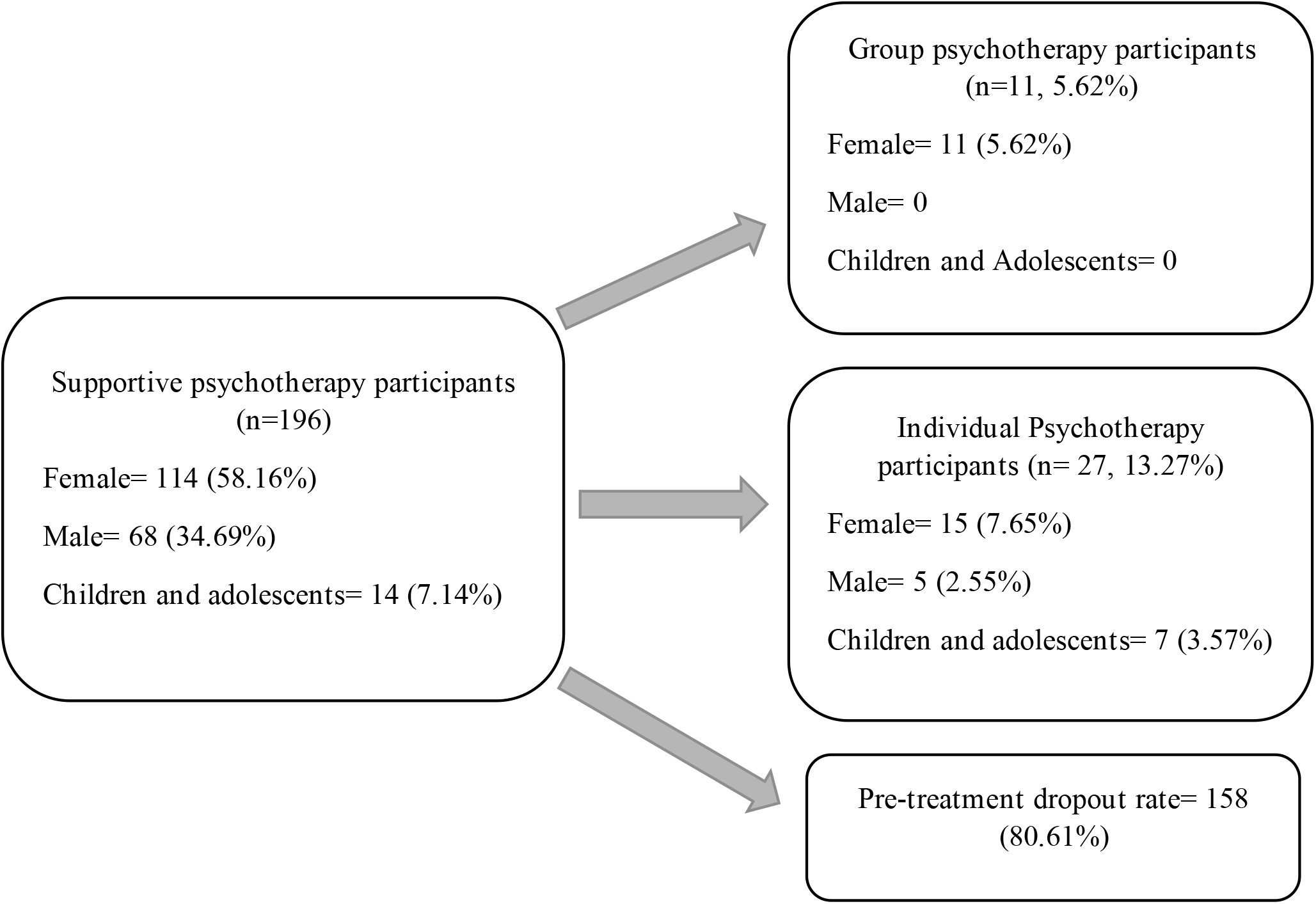
Percentage of dropouts in the supportive psychotherapy groups

#### A: Online Supportive Group Therapy

At last, only two groups of women were formed (Mean age: 40.09). One group consisted of five, and another consisted of six members. Efforts to establish men’s group therapy were unsuccessful. The average participation of members was 3.90 sessions in both established groups.

#### B: Online Individual Supportive Psychotherapy

Finally, 20 adults (Mean age: 42.85) entered individual supportive psychotherapy, and 87 individual sessions were held. The average number of sessions attended by participants was 4.23.

Regarding children and adolescents, among 14 cases who needed psychotherapy, only half of them (six girls and one boy, mean age: 9.42) attended their sessions. Most of these sessions were held as one-session psycho-education (the average number of sessions was 1.57).

## 4 Discussion

This study has shed light on the potential establishment of a support system for bereaved individuals who experienced grief during the first and second peaks of the COVID-19 pandemic in Iran. There are significant challenges we encountered and lessons learned from the study, which have been highlighted bellow:

### 4.1. Optimal Time of Initiating Intervention

The optimal timing for starting psychological interventions for grief has not been definitively determined, and a review article has reported that very early interventions do not significantly impact. However, Menichetti et al.’s (2021) study, conducted during the COVID-19 pandemic, showed that the first phone call was made within 2-3 days of the loss. In our service, contact with the bereaved was established two weeks after the death. However, because of pandemic circumstances, there were significant delays in starting contact in some cases, extending over eight weeks.

### 4.2. Effective Method of Intervention

Multiple studies have demonstrated the effectiveness of short-term supportive group therapy for bereaved adults (Hoffmann et al., 2018; Robinson and Pond, 2019; Supiano et al., 2017; Yoo and Kang, 2006). The effectiveness of such groups has been attributed to providing a place for sharing feelings, receiving emotional support, finding validation, and deriving new meaning from the traumatic experience of bereavement. Therefore, following the initial evaluation, in our study participants were encouraged to engage in group therapy. However, despite initial willingness expressed during screening calls, many participants did not adhere to group therapy or opted for individual sessions during follow-up. Similar findings were observed in the study by Shechtman et al. (2016), where both men and women showed a preference for individual therapy as they claimed that participating in group therapy raises fears of self-disclosure, judgment, and rejection more than individual therapy.

### 4.3. Pre-treatment Dropout

In our study a considerable number of dropouts were observed at the starting point; before the commencement of psychiatric visits or supportive psychotherapy. Studies have indicated that more than half of these dropouts occur during the first session or early in the treatment (Thormählen et al., 2003). In general, dropouts in psychological treatments are a common issue, with reported rates between 30% and 60% (Reis and Brown, 1999) but our system experienced a higher rate with 74.58% in psychiatry consultation and 80.61% in supportive psychotherapy interventions. Although it is unclear which factors have played the most significant roles in this massive attrition, various obstacles could be considered potential barriers. Above all, we assume that the unique nature of COVID-19, the absence of definitive treatments, particularly during the early stages of the pandemic, the complex grief process, the inability to visit the deceased in their final days, and the suddenness of death can trigger various defense mechanisms in survivors, leading to dissatisfaction with interventions and difficulties in establishing a good therapeutic alliance. The delay in making initial contact in our service was another factor that led to a higher dropout rate, as some survivors had already received treatment from therapists or their grief symptoms had resolved over time.

### 4.4. Technical Difficulties

Technical problems, such as the lack of access to smartphones, difficulties with software installation and usage, and poor internet connectivity, were identified in various studies as barriers to treatment, particularly in the context of group therapy. For instance, inadequate utilization of the program resulted from issues such as the inability to control microphone and camera settings and the absence of a private and quiet space for uninterrupted participation, could negatively impact the establishment of a therapeutic alliance and intra-session processes. Efforts were made to address these problems by providing instructions to group members before each session and reminding these instructions at the beginning of each session, but these issues persisted throughout the study. Consequently, the low average participation in group therapy may indicate a failure in establishing a proper therapeutic alliance.

To mitigate technical issues, individual supportive psychotherapy sessions were conducted via voice calls for participants unable to engage in online video sessions. Furthermore, the waiting time was reduced by initiating sessions immediately after the assessment. However, the system still experienced a relatively high dropout rate, suggesting that technical problems and waiting times were not the primary reasons for attrition. A lack of trust in the treatment system and the inability to establish a therapeutic alliance may serve as significant barriers to seeking help. Menichetti et al. also emphasized the unique characteristics of these psychological interventions, including the absence of a typical setting, not face-to-face contact, multiple and somewhat ambiguous aims, and interactions during a sensitive period, which can contribute to the challenges in engaging and retaining participants.

### 4.5. Gender-specific Participation

More than 78% of those who attended psychiatry visits and over 84% of those who underwent psychotherapy were women, highlighting a disparity in our service system between men and women in expressing the need for mental health services and receiving them. Various studies have shown that women refer to both physical and mental health services more than men (Brown et al., 2019; Thompson et al., 2016). It has also been noted that men are more likely to discontinue therapy (Melville et al., 2010). Various reasons have been assumed, including the masculine role society imposes on men. Moreover, the article has also mentioned that most men facing problems try to avoid the community and, at the same time, increase working hours to adapt to the situation (Brown et al., 2019).

### 4.6. Children and Adolescents

The last issue we dealt with was the process of grief in children and adolescents. Although our primary interventions were designed for grieving adults, we also expected issues regarding grieving children. So, we included a child and adolescent psychiatrist and a child and adolescent psychologist in our service. Overall, a single psycho-educational session for the family regarding how to deal with children’s grief met the needs of the families.

## 5 Conclusion

In conclusion, while the establishment of a support system for bereaved individuals during the COVID-19 pandemic in Iran is a valuable initiative, this study highlighted several challenges. The high dropout rate, which could be attributed to a lack of trust in medical professionals and difficulties in establishing a therapeutic alliance, indicates the need for further exploration of strategies to engage and retain participants. Technical issues and delays in initiating contact also contributed to the dropout rate but were not the primary reasons for attrition. Future research and interventions should consider these challenges and focus on addressing the unique needs and concerns of bereaved individuals during times of crisis.

## Data Availability

All data produced in the present work are contained in the manuscript.

## Ethical consideration

The study protocol was approved by the ethical committee of the Tehran University of Medical Sciences (IR.TUMS.VCR.REC.1399.256). Participation in this study was voluntary. Verbal informed consent was obtained from participants.

## Acknowledgment

We would like to show our gratitude to Dr. Solmaz Alaei, Dr. Mamak Tahmasebi, Tahereh Jamaloo, Hasan Samadi, Fatemeh Bahrami, Shima Shirafkan, and Fatemeh Sabzi for their scientific contribution and assistance in conducting groups during the project. We also thank the contributions of staff members and residents of Roozbeh Psychiatric Hospital and the University of Social Welfare and Rehabilitation.

