## Supplemental Table 1 for "Online early supportive psychological intervention for bereaved families during COVID-19: Lessons Learned and Implementation Challenges"

| | Rules of group therapy |
|---|---|
| 1 | It is recommended to keep your video on while participating in the group. |
| 2 | Be sure to choose a private and safe environment to participate in the group. |
| 3 | You are not allowed to record the group meetings. |
| 4 | You are recommended to use hands-free. |
| 5 | It would help to respect other members and not interrupt others when talking. |
| 6 | One of the essential goals of therapy is to provide a suitable and safe environment for thinking and talking about feelings. Acting on these feelings, such as humiliation, insults, or shouting instead of talking about anger, could disrupt the group process. So, you are expected to talk about your feelings, not act on them. |
| 7 | You are expected to attend all the sessions. |
| 8 | You are asked to inform the therapist and other members about future absences or the decision to leave the group. |
| 9 | You are expected to join the meetings with a device other than a mobile phone and turn off your mobile phones during the meeting. |
| 10 | Any activity that disrupts the group process (such as reading, eating, etc.) should be avoided. |
| 11 | Maintaining confidentiality is one of the essential principles in group therapy. Therefore, group members should not transfer the issues raised in the group outside. |
| 12 | You preferably should not know other members of the group in advance, in order to talk about your feelings more easily. So, if you know a group member, please convey this to the therapist. |
