## Supplemental Table 2 for "Online early supportive psychological intervention for bereaved families during COVID-19: Lessons Learned and Implementation Challenges"

Dos and don’ts in working with bereaved patients

| | |
|---|---|
| 1 | Listen carefully; your presence is more helpful than you think. |
| 2 | Saying short words of condolence like “I'm so sorry" or "These are hard days" is enough. Do not use phrases like "They had a good life" or "They’re not suffering anymore." These expressions trivialize the experience of loss. |
| 3 | Never tell the clients that you understand their experience; every relationship and every grief feeling has a unique sense. |
| 4 | Use direct words such as "death"; do not encourage client denial by using indirect words. |
| 5 | Your body language must convey openness. In online meetings, it is better that you can be fully visible to the patients, and members should be asked to make visual contact as much as possible. |
| 6 | When necessary, allow the silence to continue. |
| 7 | Do not interrupt people, and don't stop the members from crying. |
| 8 | Do not give advice. |
| 9 | Never talk about your own experience of grief. Instead, keep the focus on the client's experience. |
| 10 | Answer the questions honestly. |
| 11 | Acknowledge what the client says but maintain a neutral position in the conversation. |
| 12 | Let the conversation flow on its own, do not direct it in a way you like. |
| 13 | Tell the client: "you are crying" rather than "you are sad" so that he/she can express the feelings behind his/her behavior. |
| 14 | Be careful about your body language |
| 15 | Do not look at your notes, watch, etc., while talking. |
| 16 | Ask clients, "What happened?" Most people around the grieving person avoid this question and act as if nothing happened, increasing the feeling of isolation in the grieving people. At the same time, do not burden the clients to talk about their experiences. |
| 17 | Strengthen positive coping mechanisms when necessary, such as eating and sleeping regularly, talking to family members or friends via video contact, engaging in routine activities such as exercise, walking, or praying |
| 18 | Stay away from the news of the disease and its consequences (Reynolds et al., 2007). |
